# How dietary landscapes impact food allergy

**DOI:** 10.1101/2024.04.06.24305435

**Authors:** Duan Ni, Alistair Senior, Jian Tan, Laurence Macia, Ralph Nanan

**Author notes:** **Corresponding author** Ralph Nanan, Sydney Medical School Nepean, The University of Sydney., Nepean Hospital, Level 5, South Block, Penrith NSW, 2751, Australia.

## Abstract

Diets and environments are critical determinants for food allergy development. Harnessing unprecedented epidemiological and nutritional data, we examined the overall dietary environments for common food allergens and their intrinsic nutrient composition. We found that food and macronutrient supplies minimally impacted food allergy prevalence, but higher protein and glycine in food allergens correlated with less allergies. These findings offer new directions in food allergy research and management.

## Main

Food allergies is emerging as a significant public health concern^1,2^. One of the central ambitions in allergy research is to characterize the risk factors for food allergy. Previous research mostly focused on the macroenvironments (hygiene hypothesis), the individuals’ biological characteristics, and immunological aspects with gut microbiome as an intermediary^1^. Factors that have been neglected, however, include the intrinsic properties of common trigger foods, like their nutrient compositions, and their extrinsic environments, like interactions with other dietary components.

Furthermore, early life exposure to allergenic foods is linked to lower risks of food allergies^3-5^. Nevertheless, little is known about whether similar protective effects apply to exposures to specific food environments and nutrient environments. Moreover, studies towards what food components trigger allergy have mostly concentrated on allergenic proteins recognized by the immune system^6^. However, accumulating evidence on involvements of other macronutrients, like fats and carbohydrates, in allergenic sensitization^7-9^, suggested that a more holistic overview towards food components and their potential interactions is needed.

Harnessing comprehensive epidemiological and nutritional data, we systematically analyzed the extrinsic and intrinsic nutritional factors associated with the prevalence of common food allergies. We interrogated how food and nutrient environments correlate with food allergy prevalence and how the nutrient compositions of foods are linked to allergenicity. Survey of a collection of food allergy studies for the 8 most common food allergens found that extrinsic dietary environments, reflected by the food and macronutrient supplies, minimally impacted food allergy prevalence. However, a robust association was observed between the intrinsic protein and glycine content in food allergens and reduced allergy prevalence.

We harnessed a dataset investigating allergies for the 8 common food allergens (milk, egg, peanut, tree nut, wheat, soy, fish and shellfish) in Europe from 2001 to 2021^10^ (Figure 1A). This dataset presented a robust epidemiological landscape in a broad spatiotemporal context for these common and well-characterized food allergies. Figure 1B summarized the pooled estimates for all age self-reported food allergy lifetime and point prevalence. Milk and egg exhibited the highest lifetime and point prevalence respectively.

**Figure 1.**
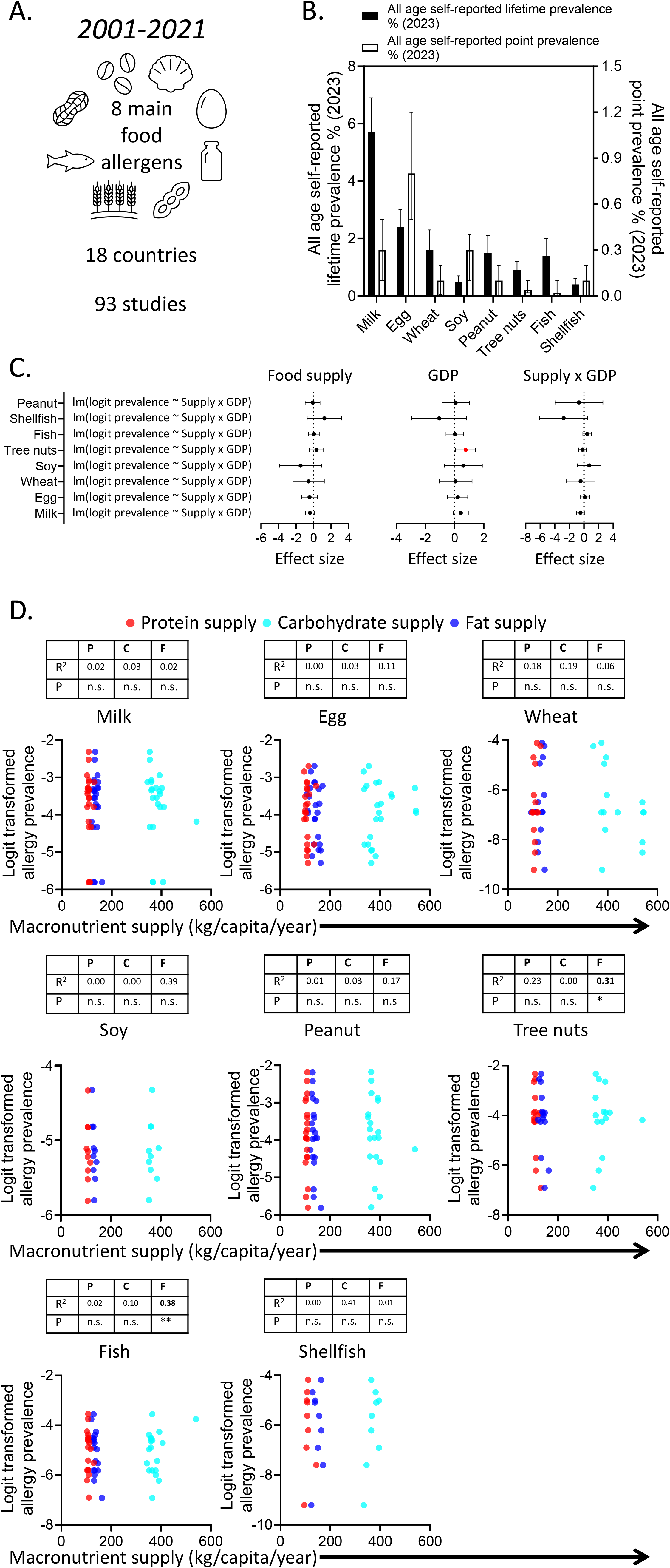
**A-B**. Overview of allergy studies for the 8 common food allergens (milk, egg, wheat, soy, peanut, tree nut, fish and shellfish, **A**) and their pooled results (**B**). **C**. Effect sizes of the linear modellings for scaled food allergen supplies (kg/capita/year), scaled GDP per capita (U.S. dollars) and the corresponding logit transformed food allergy prevalence. Error bars depict 95% confidence intervals. Red dot denotes statistical significance. **D**. Correlation analyses for the logit transformed food allergy prevalence and their corresponding national protein (P, red), carbohydrate (C, cyan) and fat (F, blue) supplies (kg/capita/year) of the same year.

Prevalence data were logit transformed and the food and macronutrient data of the same year were collated for analysis. Annual gross domestic product (GDP) was included for adjustments. GDP reflects the socioeconomic status/wealth, potentially influencing allergy prevalences^2^.

Early introduction of allergenic foods confers protection against food allergies later in life^3-5^. To test whether exposures via dietary environments confers similar effects, food supplies were used as proxies, and modelled against corresponding food allergy prevalence. Linear models were fitted for the food allergy prevalence against the corresponding scaled food supply and GDP data, with consideration of their potential interactions. As shown in Figure 1C, there was no significant effect of the supplies of the 8 individual allergens on allergy prevalence. Only for tree nut, GDP showed a significant effect. These results suggested that exposures via allergenic food supplies and the socioeconomic environments, were not generally associated with corresponding food allergy prevalence.

We previously showed that allergic diseases could be modulated via diets and were associated with the overall nutrient environments, another critical aspect of dietary exposures^11-13^. Food allergy prevalence was thus compared against the macronutrient supplies of the corresponding countries and time points. We found no association, except for tree nut and fish exhibiting negative correlations with fat supplies (Figure 1D). This indicated that the overall nutrient environments, represented by macronutrient supplies, minimally influenced food allergy prevalence.

After extrinsic environments, we next attempted to characterize the intrinsic properties of food allergens, examining not only proteins, the main focus previously, but also other nutrient components and their potential interactions.

Macronutrient compositions of the 8 common food allergens were calculated (Figure 2A-B) and compared against the lifetime food allergy prevalence estimates in 2014^14^ and 2023^10^. As shown in Figure 2C-D, protein compositions in foods were negatively correlated with their corresponding food allergy prevalence, while no association was found for other components (Figure S1A-C). Analysis based on individual studies yielded similar result (Figure 2E). To account for potential random confounding effects from specific foods or the countries where the studies were carried out, an array of linear mixed-effect models were fitted. Nutrient compositions of the potential allergenic foods were used as predictors. Foods and countries the data was based on were adjusted as random effects. A model considering the effects from protein content with additive random effects from foods and countries was favoured based on Akaike information criterion (Figure 2F, Table S1). It also unveiled a robust negative correlation between protein content and allergy prevalence (Figure 2G). Significantly, these associations were validated in independent American and Canadian datasets^15^ (Figure S1D-G).

**Figure 2.**
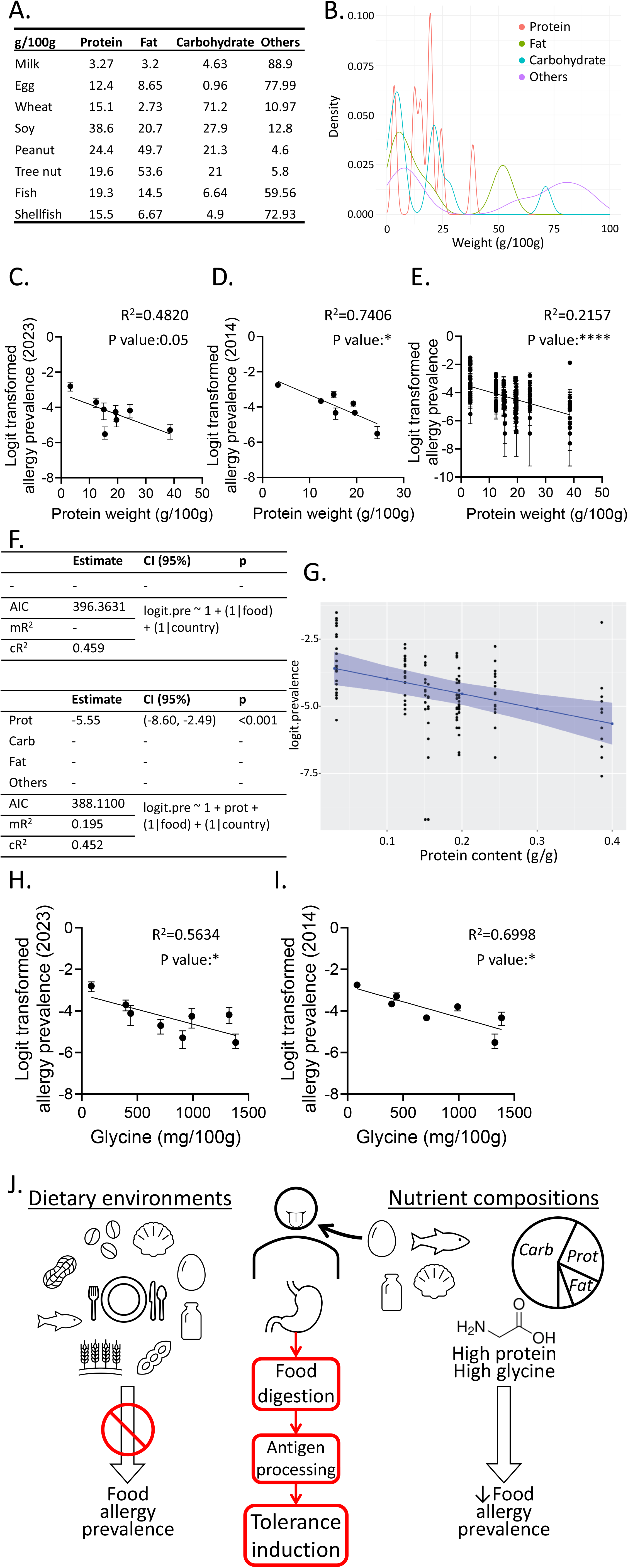
**A-B**. Overview of the nutrient compositions (g content/100g food, **A**) and distributions (**B**) of the 8 common food allergens. **C-D**. Correlation analyses for the protein content in food allergens and their corresponding logit transformed allergy prevalence estimates in 2023 (**C**) and 2014 (**D**). **E**. Correlation analysis for the protein content in food allergens and their corresponding logit transformed allergy prevalence reported in 93 studies. **F**. Statistical outputs for the null model and the chosen linear mixed-effect model analyzing protein content in food allergens and their corresponding logit transformed allergy prevalence. **G**. Estimate plot for the effect of protein content (g/g food) in food allergens and their corresponding logit transformed allergy prevalence based on the chosen linear mixed-effect model. **H-I**. Correlation analyses for the glycine content (mg/100g food) in food allergens and their corresponding logit transformed allergy prevalence estimates in 2023 (**H**) and 2014 (**I**). **J**. Model for the impacts of dietary environments and nutrient compositions on food allergy.

We further interrogated whether amino acids might exert similar effects. Indeed, glycine content exhibited consistent negative correlations with allergy prevalence (Figure 2H-I).

Together, our analyses revealed that protein and glycine content, in the 8 common food allergens, negatively correlated with their allergy prevalence.

Leveraging epidemiological and nutritional data, we systematically interrogated how extrinsic and intrinsic nutritional factors are linked to food allergy prevalences. While dietary environments lacked evident effects, higher protein and glycine content in food allergens correlated with lower allergy prevalence.

It is established that early life exposure to food allergens induces tolerance against allergies^3-5^. Our results suggested that exposures reflected by supplies and availabilities of allergenic foods and dietary macronutrients did not confer such protective effect. This warrants further investigations regarding the threshold and timing of exposures for allergy protection.

Instead of focusing solely on food allergenic proteins, our analyses delineated the contributions to allergenicity from different nutrient components and their plausible interactions, shifting the paradigm for interrogating the intrinsic determinants of food allergenicity. We found that higher protein content in trigger foods was associated with lower allergy prevalence, while other nutrient components conferred negligible influences. Specifically, glycine content was linked to reduced food allergy prevalence. Our findings were based on the 8 most common food allergens, with well-characterized clinical features and well-documented epidemiological data. Further high quality epidemiological and nutritional data is needed to validate our discoveries beyond aforementioned 8 allergens. Additionally, similar findings were found for both European and North American populations. It remains to be tested if such results would pertain at a broader global scale.

It is unclear how protein content modifies food allergenicity (Figure 2J). High protein content might alter the food digestibility, interfering with allergen exposure to the host and thus tolerance induction. In this context, the amounts of proteins in allergenic foods are likely to also affect antigen uptake and processing, hence influencing allergenic sensitization.

Apart from quantity, protein qualitative properties of trigger foods might be equally important. Glycine content in food allergens correlated with decreased allergy prevalence. Its small size might tweak peptide flexibility and further protein biophysical features. This might also interfere with protein digestion, and/or antigen processing and presentation. Additionally, glycine independently exhibits potent immunomodulatory capacities^16^, suppressing acute allergic responses^17^. Whether these properties could extend to high glycine foods remains to be uncovered.

Together, our study sheds unprecedented insights towards dietary factors influencing food allergy by comprehensive surveying their epidemiological and nutritional landscapes, opening novel avenues for allergy research. Our findings prompt future studies to unravel the mechanistic and pathophysiology of food allergies in a broader dietary context, which could guide food allergy managements and/or prevention.

## Methods

Food allergy data and nutritional data were collected as described in Supplementary Information. Analyses were run in RStudio (v4.1.2) with *stats* and *lme4* packages. Detailed methods are available in Supplementary Information.

## Data Availability

All data used in the present study are publicly available

## Data availability

All data used in the present study are publicly available as described in Supplementary Information.

## Acknowledgements

This project is supported by the Norman Ernest Bequest Fund.

## Contributions

DN participated to the study design, performed most of the analyses and wrote the manuscript. AS, JT, and LM participated in the analyses and data interpretation. RN supervised the study and wrote the manuscript.

All authors read and approved the final manuscript.

## Ethics declarations

LM is a current employee of the Translational Science Hub Global Sanofi Vaccines R&D Brisbane, Australia. Her contribution to this work was when she was an employee of the University of Sydney. The other authors declare no competing interests.

## References

1 Renz, H. et al. Food allergy. Nat Rev Dis Primers 4, 17098, doi:10.1038/nrdp.2017.98 (2018).

2 Shin, Y. H. et al. Global, regional, and national burden of allergic disorders and their risk factors in 204 countries and territories, from 1990 to 2019: A systematic analysis for the Global Burden of Disease Study 2019. Allergy 78, 2232–2254, doi:10.1111/all.15807 (2023).

3 Trogen, B., Jacobs, S. & Nowak-Wegrzyn, A. Early Introduction of Allergenic Foods and the Prevention of Food Allergy. Nutrients 14, doi:10.3390/nu14132565 (2022).

4 Frazier, A. L., Camargo, C. A., Jr., Malspeis, S., Willett, W. C. & Young, M. C. Prospective study of peripregnancy consumption of peanuts or tree nuts by mothers and the risk of peanut or tree nut allergy in their offspring. JAMA Pediatr 168, 156–162, doi:10.1001/jamapediatrics.2013.4139 (2014).

5 Bunyavanich, S. et al. Peanut, milk, and wheat intake during pregnancy is associated with reduced allergy and asthma in children. J Allergy Clin Immunol 133, 1373–1382, doi:10.1016/j.jaci.2013.11.040 (2014).

6 Breiteneder, H. & Mills, E. N. Molecular properties of food allergens. J Allergy Clin Immunol 115, 14–23; quiz 24, doi:10.1016/j.jaci.2004.10.022 (2005).

7 Bublin, M., Eiwegger, T. & Breiteneder, H. Do lipids influence the allergic sensitization process? J Allergy Clin Immunol 134, 521–529, doi:10.1016/j.jaci.2014.04.015 (2014).

8 Del Moral, M. G. & Martinez-Naves, E. The Role of Lipids in Development of Allergic Responses. Immune Netw 17, 133–143, doi:10.4110/in.2017.17.3.133 (2017).

9 Soh, J. Y., Huang, C. H. & Lee, B. W. Carbohydrates as food allergens. Asia Pac Allergy 5, 17–24, doi:10.5415/apallergy.2015.5.1.17 (2015).

10 Spolidoro, G. C. I. et al. Prevalence estimates of eight big food allergies in Europe: Updated systematic review and meta-analysis. Allergy 78, 2361–2417, doi:10.1111/all.15801 (2023).

11 Ni, D. et al. Global associations of macronutrient supply and asthma disease burden. Allergy, doi:10.1111/all.16067 (2024).

12 Tan, J. et al. Dietary Fiber and Bacterial SCFA Enhance Oral Tolerance and Protect against Food Allergy through Diverse Cellular Pathways. Cell Rep 15, 2809–2824, doi:10.1016/j.celrep.2016.05.047 (2016).

13 McKenzie, C., Tan, J., Macia, L. & Mackay, C. R. The nutrition-gut microbiome-physiology axis and allergic diseases. Immunol Rev 278, 277–295, doi:10.1111/imr.12556 (2017).

14 Nwaru, B. I. et al. Prevalence of common food allergies in Europe: a systematic review and meta-analysis. Allergy 69, 992–1007, doi:10.1111/all.12423 (2014).

15 Messina, M. & Venter, C. Recent Surveys on Food Allergy Prevalence. Nutrition Today 55, 22–29, doi:10.1097/nt.0000000000000389 (2020).

16 Aguayo-Ceron, K. A. et al. Glycine: The Smallest Anti-Inflammatory Micronutrient. Int J Mol Sci 24, doi:10.3390/ijms241411236 (2023).

17 van Bergenhenegouwen, J. et al. Oral exposure to the free amino acid glycine inhibits the acute allergic response in a model of cow’s milk allergy in mice. Nutr Res 58, 95–105, doi:10.1016/j.nutres.2018.07.005 (2018).

