## Supplementary Information for "How dietary landscapes impact food allergy"

**Supplementary Materials and Methods**

**Data collection and processing**

Food allergy prevalence data were curated from the original studies^1,2^ and were logit transformed for analysis. Food and macronutrient supply data were collated from the Food and Agriculture Organization of the United Nations (FAOSTAT) database (<https://www.fao.org/faostat/en/#data>). Annual gross domestic product (GDP) per capita data were extracted from the Maddison project^3^. Nutritional compositions of food allergens were obtained from the U.S. Department of Agriculture (USDA) FoodData Central database (<https://fdc.nal.usda.gov/index.html>). Each macronutrient (protein, carbohydrate and fat) was calculated as g content per 100g of food and the remaining was grouped as other.

**Modelling and analysis**

Data analyses were run in RStudio (v4.1.2.) or in GraphPad PRISM.

*lm()* function in *stats* package was used for the linear modelling of logit transformed food allergy prevalence and the corresponding annual food allergen supply of the year when the studies were carried out and the country GDP of the same time point. Food supply and GDP data were first scaled, and their interactions were accounted within the analysis.

GraphPad PRISM was used for the correlation analyses for logit transformed food allergy prevalence and the corresponding annual macronutrient (protein, carbohydrate and fat) supplies.

GraphPad PRISM was also used for the correlation analyses for logit transformed food allergy prevalence and the corresponding macronutrient content and amino acid content.

*lmer()* function in *lme4* package was used for the linear mixed-effects modelling of logit transformed food allergy prevalence and the corresponding macronutrient compositions, with foods and countries that the prevalence data was based on adjusted as random effects. The macronutrient compositions and their potential interactions were used as predictors, and a null model considering only the random effects from foods and countries was included as well. Akaike information criterion (AIC) was used for model evaluation and the one with the lowest AIC was favoured. If the differences of the AIC results of two models were less than 1, the simpler model was favoured.

**Supplementary Figure**

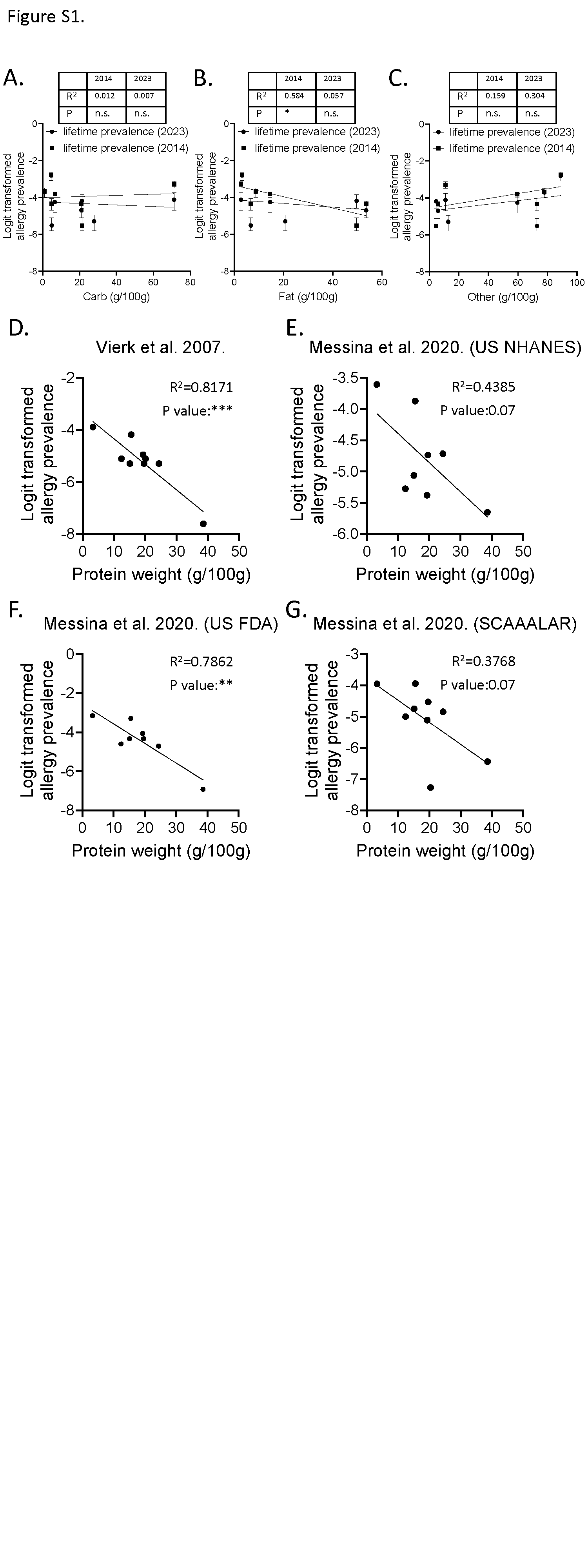

**Figure S1. A-C.** Correlation analyses for the carbohydrate (carb, **A**), fat (**B**), and other (**C**) contents in food allergens and their corresponding logit transformed allergy prevalence. **D-G.** Correlation analyses for the protein content in food allergens and their corresponding logit transformed allergy prevalence in study by Vierk et al. (**D**), and study by Messina et al. (**E-G**). (NHANES, National Health and Nutrition Examination Survey; FDA, Food and Drug Administration; SCAAALAR, Surveying Canadians to Assess the Prevalence of Food Allergies and Attitudes Towards Food Labelling and Risk)

**Supplementary Table**

**Table S1.** Akaike information criterion (AIC) results for the linear mixed-effect modelling for logit transformed food allergy prevalence (logit.pre) and the corresponding nutrient compositions (protein, carbohydrate (carb), fat, and other), with the corresponding foods and countries that the data was based on adjusted as random effects.

| **Models** | **AIC** |
| --- | --- |
| lmer(logit.pre ~ 1+ (1\|Food) + (1\|Country)) | 435.6623 |
| lmer(logit.pre ~ 1+ (1\|Food)) | 445.2116 |
| lmer(logit.pre ~ 1+ (1\|Country)) | 467.2911 |
| **lmer(logit.pre ~ 1+ Protein + (1\|Food) + (1\|Country))** | **429.8765** |
| lmer(logit.pre ~ 1+ Protein + (1\|Food)) | 439.8596 |
| lmer(logit.pre ~ 1+ Protein + (1\|Country)) | 437.7819 |
| lmer(logit.pre ~ 1+ Fat + (1\|Food) + (1\|Country)) | 434.919 |
| lmer(logit.pre ~ 1+ Fat + (1\|Food)) | 444.3684 |
| lmer(logit.pre ~ 1+ Fat + (1\|Country)) | 463.3676 |
| lmer(logit.pre ~ 1+ Carb + (1\|Food) + (1\|Country)) | 434.7021 |
| lmer(logit.pre ~ 1+ Carb + (1\|Food)) | 444.5159 |
| lmer(logit.pre ~ 1+ Carb + (1\|Country)) | 460.8824 |
| lmer(logit.pre ~ 1+ Other + (1\|Food) + (1\|Country)) | 434.7567 |
| lmer(logit.pre ~ 1+ Other + (1\|Food)) | 444.4665 |
| lmer(logit.pre ~ 1+ Other + (1\|Country)) | 451.2173 |
| lmer(logit.pre ~ 0+ Protein + Fat + Carb + Other + (1\|Food) + (1\|Country)) | 429.0321 |
| lmer(logit.pre ~ 0+ Protein + Fat + Carb + Other + (1\|Food)) | 439.1997 |
| lmer(logit.pre ~ 0+ Protein + Fat + Carb + Other + (1\|Country)) | 438.4033 |
| lmer(logit.pre ~ 1+ Protein + Fat + Carb + (1\|Food) + (1\|Country)) | 429.0321 |
| lmer(logit.pre ~ 1+ Protein + Fat + Other + (1\|Food) + (1\|Country)) | 429.0321 |
| lmer(logit.pre ~ 1+ Protein + Carb + Other + (1\|Food) + (1\|Country)) | 429.0321 |
| lmer(logit.pre ~ 1+ Fat + Carb + Other + (1\|Food) + (1\|Country)) | 429.0321 |

**References**

1	Spolidoro, G. C. I. *et al.* Prevalence estimates of eight big food allergies in Europe: Updated systematic review and meta-analysis. *Allergy* **78**, 2361-2417, doi:10.1111/all.15801 (2023).

2	Nwaru, B. I. *et al.* Prevalence of common food allergies in Europe: a systematic review and meta-analysis. *Allergy* **69**, 992-1007, doi:10.1111/all.12423 (2014).

3	Jutta Bolt, R. I., Herman de Jong, and Jan Luiten van Zanden. Vol. GD-174	(Groningen Growth and Development Center, GGDC Research Memorandum, 2018).
